# Automated measurement of the foveal avascular zone in healthy eyes on Heidelberg Spectralis Optical Coherence Tomography Angiography

**DOI:** 10.1101/2020.08.06.20169672

**Authors:** Laura Gutierrez-Benitez, Yolanda Palomino, Noe Casas, Mouafk Asaad

## Abstract

**Purpose:** To develop and evaluate an automated method to measure the foveal avascular zone (FAZ) area in healthy eyes on Heidelberg Spectralis Optical Coherence Tomography Angiography (HS-OCTA). This method is referred to as the modified Kanno-Saitama macro (mKSM) and it is an evolution of the original Kanno-Saitama macro (KSM) approach.

**Methods:** This cross-sectional study included 29 eyes of 25 healthy volunteers who underwent HS-OCTA at the macular area twice at the same time. Regardless of the quality of the images, all of them were included. Macular data on the superficial vascular plexus, intermediate capillary plexus and deep capillary plexus were processed by mKSM. The FAZ area was measured twice automatically using the mKSM and KSM and twice manually by two independent examiners.

**Results:** From 174 images, KSM could not measure correctly 31% while mKSM could successfully measure all of them. Intrascan intraclass coefficient ranged from 0,948 to 0,993 for manual measurements and was 1 for mKSM method, which means that mKSM FAZ area value is always the same for the same OCTA image. Despite that the difference between human examiners is smaller than between human examiners and mKSM according to Bland-Altman plots, the scatterplots show a strong correlation between human and automatic measurements. The best results are obtained in intermediate capillary plexus.

**Conclusions:** With mKSM, the automated determination of the FAZ area in HS-OCTA is feasible and less human-dependent. It solves the inability of KSM to measure the FAZ area in suboptimal quality images which are frequent in daily clinical practice. Therefore, the mKSM processing could contribute to our understanding of the three vascular plexuses.

## INTRODUCTION

Optical coherence tomography angiography (OCTA) has been a significant advance for the visualization of retinal vascularization, without being an invasive test, since it does not require intravenous dye injection, offering results comparable to those obtained with fluorescein angiography (1)(2)(3)(4). The use of OCTA provides high-quality images that can be obtained quickly and reproducibly. It enables the evaluation of multiple retinal pathologies in clinical practice, particularly underlying macular vascular diseases affecting the foveal avascular zone (FAZ) (5).

Earlier studies have reported that the FAZ area size varies significantly among healthy eyes. Although there are controversial results among the studies, the FAZ area has been found to increase with age (6) (7), to be larger in women (8)(7), smaller when increased axial length, thicker central retinal thickness or increased vascular density (9) (10) (11) (12). It has also been found to be larger in asian people than in caucasians(13) and larger in caucasians than in black people (14). Besides, glaucoma with central visual field defects (15)(16)(17) and several ischemic pathologies such as diabetic retinopathy and retinal venous occlusion (18)(19) have been reported to cause an enlarged FAZ area.

The OCTA FAZ measurement techniques have advanced in recent years (20) evaluating the characteristics of the FAZ area, either in healthy patients or in different pathologies. The main types of OCTA that are commonly used today are spectral-domain OCTA (SD-OCTA) and swept-source OCTA (SS-OCTA). Among them, there are OCTA devices that incorporate embedded software that automatically measures the FAZ area, such as AngioVue OCTA (Optovue, Inc, Fremont, CA) or the SS-OCTA (Plex Elite 9000, Carl Zeiss Meditec Dublin, CA) (21)(22). Some other devices, such as Heidelberg Spectralis OCTA (HS-OCTA), do not incorporate this kind of software. Therefore, the FAZ area has to be measured manually in HS-OCTA (23).

Traditionally, the FAZ area was measured on the fluorescein angiography images where it was not possible to separate the different vascular plexuses. Nowadays, the OCTA enables 3D imaging of retinochoroidal vasculature with high spatial resolution, and the different plexus can be segmented. Therefore, HS-OCTA shows the microvasculature of three distinct retinal vascular layers where the FAZ area can be measured independently on the superficial vascular plexus (SVP), the intermediate capillary plexus (ICP) and the deep capillary plexus (DCP). These microvascular layers correlate with anatomical structures(24).

The objective of our study is to develop and evaluate an automated method to measure the SVP, ICP and DCP FAZ area in HS-OCTA images, which provides objectivity, reproducibility, speed in measurements and is tolerant with images not perfectly centered or with artefacts. The automated measurements are compared to manual measurements made by two examiners, and also to other automated device-agnostic measurement method described in the literature.

The proposed automated measurement method is tolerant to images of suboptimal quality, which is a common situation in daily practice. To our knowledge, there are no previous studies that evaluate an automated method for FAZ area measurements in images acquired by HS-OCTA that offers such robustness, being common for previous studies to exclude the images with suboptimal quality, therefore biasing their evaluations.

## METHODS

### SUBJECTS

The current study is a cross-sectional study. Healthy patients were recruited from Consorci Sanitari de Terrassa (Terrassa, Barcelona, Spain) from November 2019 to January 2020. The study was conducted following the ethical standards stated in the 1964 Declaration of Helsinki and approved by the Consorci Sanitari de Terrassa Research Ethics Committee.

Subjects were included if they were 18 years or older, not affected by any ophthalmological pathology, with best-corrected visual acuity of 20/20 or better and signed an informed consent form between November 2019 and January 2020. The exclusion criteria were as follows: any cardiovascular risk factors such as diabetes mellitus, arterial hypertension, cardiovascular disease or dyslipidaemia, any clinical evidence of systemic or ocular diseases, refractive error higher than +3.0 diopters or less than −6.0 diopters, axial length exceeding 26 mm, any previous laser treatment, cataract surgery during the previous three months, and media opacities precluding adequate fundus imaging. Smoking history was recorded, but was not an exclusion criterion.

### EYE EXAMINATIONS

All subjects underwent a thorough ophthalmic examination that included best-corrected visual acuity (BCVA), slit-lamp biomicroscopy and intraocular pressure measurement by Goldman contact tonometry. Axial length was measured with optical biometer IOL Master 500 (Carl Zeiss Meditec, Germany). Optical coherence tomography and OCTA images were performed with enhanced depth imaging methods for all patients using Heidelberg Spectralis optical coherence tomography (Software Version 6.12.1, Heidelberg Engineering, Heidelberg, Germany).

The subjects were asked to choose the eye to examine first and whether they wanted both eyes examined or not.

### OPTICAL COHERENCE TOMOGRAPHY ANGIOGRAPHY

A 10° x 10° (2,9 x 2,9mm) OCTA image centered on the fovea was scanned using HS-OCTA (Software Version 6.12.1, Heidelberg Engineering, Heidelberg, Germany). En face images were acquired in High-Resolution mode, which offers a lateral resolution of 5.7 microns per pixel for the visualization of capillaries and an axial resolution of 3,9 microns per pixel that enables precise multilayer segmentation and visualization of the retinal vascular plexuses.

The HS-OCTA software generates en face images from slabs at different layers by automated segmentation, following the slab definitions proposed by Campbell et al. (25). Therefore, the superficial vascular plexus (SVP) slab is segmented as the retinal nerve fibre layer to the inner plexiform layer. The intermediate capillary plexus (ICP) is segmented between the outer 20% of the ganglion cell complex to the inner 50% of the inner nuclear layer (INL). The deep capillary plexus (DCP) is segmented between the outer 50% of the INL to the outer plexiform layer.

### IMAGE PROCESSING

Our proposed automatic foveal area estimation method relies on two previous works. The first work on which our method relies is the processing tool AngioTool software version 0.6a (02.18.14) (26), which automates the processing of OCTA images, detecting and enhancing the vascular network.

The second work on which our method relies is a study by Ishi et al. (27), where the authors proposed and evaluated a macro-based automated analysis method called Kanno-Saitama Macro (KSM) to measure the FAZ area using ImageJ software. The study concluded that this automated method was comparable to manual measurement, saving time and being less user-dependent. The KSM method is specific to the SS-OCTA equipment (Plex Elite 9000, Version 1.6.0.21130; Carl Zeiss Meditec Dublin, CA).

Our work, referred to as modified KSM or mKSM from now on, improves over KSM on the tolerance to visual artefacts and uncentered images and extends its applicability to the HS-OCTA equipment. For that, mKSM first takes an OCTA image and applies the processing from AngioTool, obtaining a cleaner version of the vascular network image. Then, mKSM estimates the center of the foveal area by computing the center of mass of the image pixels, taking the pixel brightness value as its mass. Detecting the center of the foveal area instead of assuming that it is located at the center of the image, grants mKSM tolerance to uncentered OCTA images. Once the center of the foveal area has been estimated, mKSM applies a random 2D shift to it and computes the foveal area by means of the KSM approach. This measure is repeated 20 times, with different values for the random 2D shift of the foveal area center. The final area is computed by taking the median value of all the previous measures (to deterministically obtaining the same result for an input OCTA image, we use a fixed sequence of random numbers) (see figure 1). This area sampling process, together with the median, grants mKSM tolerance to visual artefacts in the foveal area. In KSM, such artefacts were misidentified for the fovea itself. The median of the area sample allows discarding such misidentifications because they result in outliers in the estimated area distribution. There are two problems of KSM that are solved by mKSM: misidentifications of the FAZ area due to visual artefacts and excessive area attribution to the fovea, due to low contrast of the vascularized area around the fovea. We label these problems respectively as “misidenfication” and “overmeasurement” (see figure 2). mKSM is implemented as an extension to the scientific imaging tool ImageJ.

**Figure 1.**
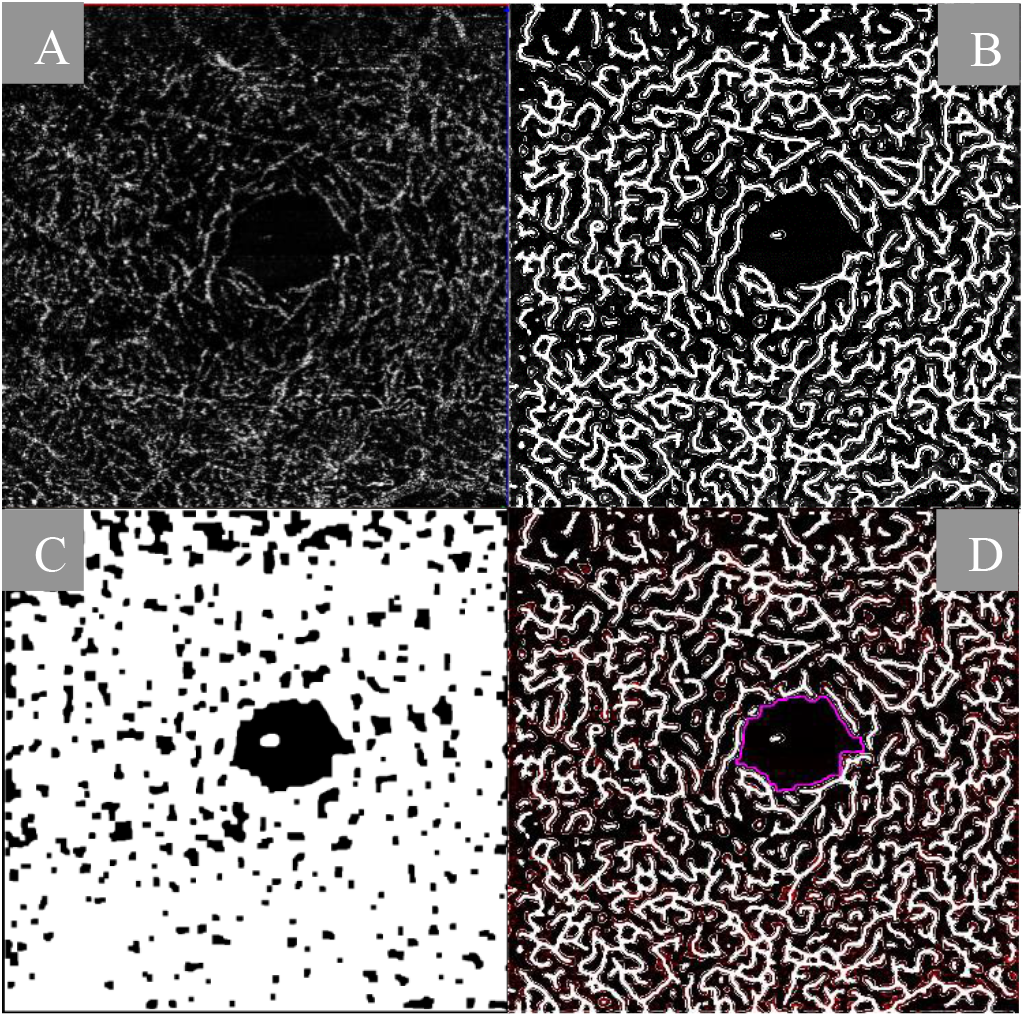
Optical coherence tomography angiography scan (A) processed by angioTool (B) and by mKSM (C). The foveal avascular zone area is drawn pink on the angioTool image (E).

**Figure 2.**
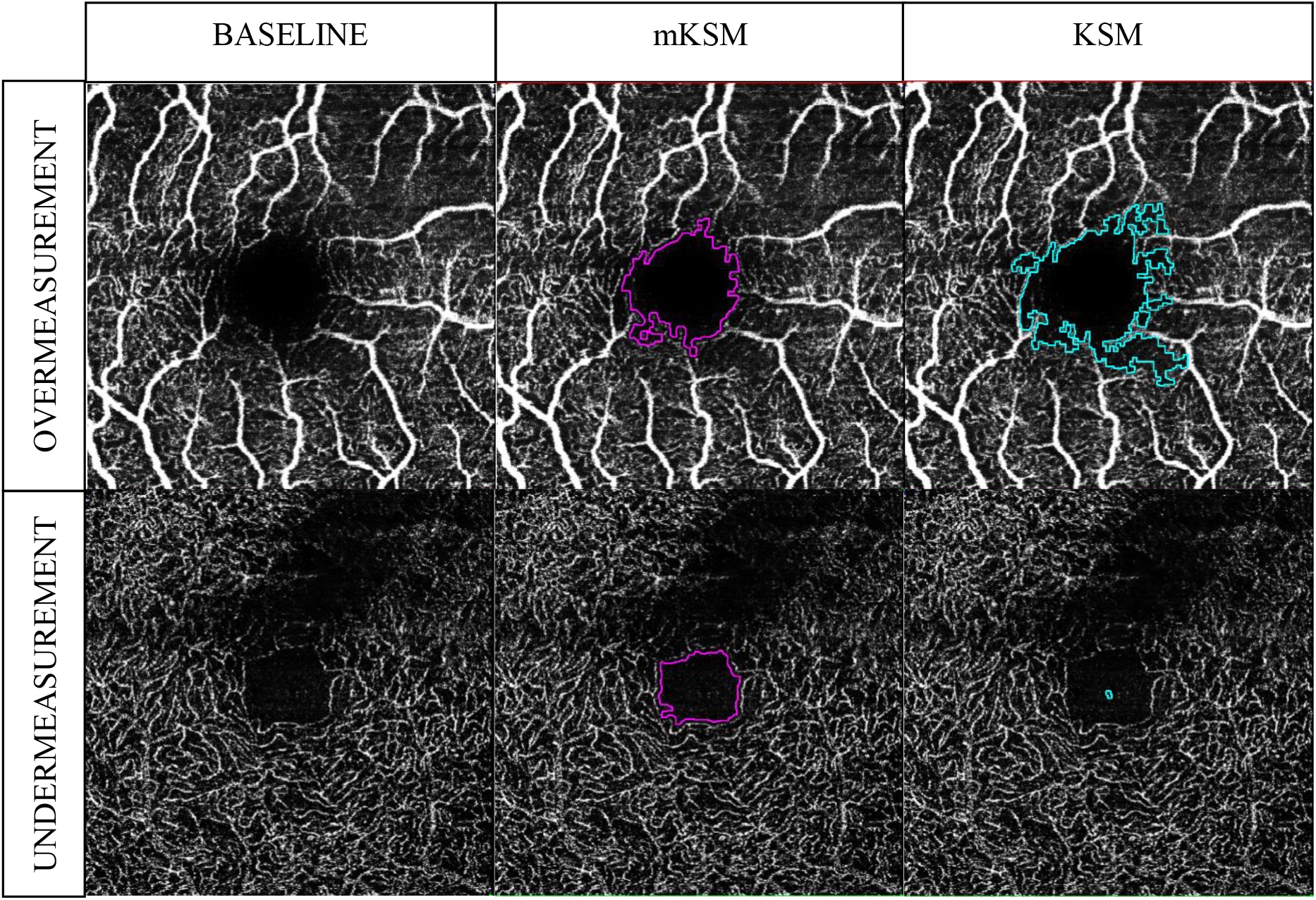
Examples of how a suboptimal image quality affects the localization of the FAZ area in other methods (KSM), while ours (mKSM) is robust and identifies it correctly: 1st column shows the original optical coherence tomography angiography scans with artefacts, 2nd column shows the foveal avascular foveal zone (FAZ) area measured by our method (mKSM), 3rd column shows the measure by other methods (KSM), which suffer from overmeasurement of the FAZ area in the first row and from a misidentification of the FAZ area in the second row.

### MANUAL MEASUREMENTS

The OCTA images were imported in the scientific image processing tool ImageJ. The scale parameter of the software was set to define a 964-pixel width in the images as 2,96mm. The manual measurements were performed by two masked examiners (YP and LG) who independently drew the FAZ area contour joining the internal points of the vessels on all the images using the polygon selections tool. The FAZ area was measured in square millimeters.

### TESTING PROTOCOL

All subjects were scanned in the morning twice on the same day at the same time. All the images were measured twice manually by both examiners, and automatically by KSM and mKSM methods to calculate the intrascan reproducibility. Interscan reproducibility was calculated using the first measurements obtained in the first and second scans for both examiners and mKSM method. The mKSM measurements were compared with manual and KSM measurements.

### STATISTICAL ANALYSES

IBM® SPSS® Statistics 25.0 and MedCalc Version 19.3.1 are used for all statistical analyses. The data for quantitative variables are expressed as the mean value and standard deviation (SD). The data for qualitative variables are expressed as an absolute number and relative frequency.

Both the the intrascan reproducibility (similarity between the measurements of the same image with the same measurement method, e.g. mKSM, manual) and the interscan reproducibility (similarity between the measurements of two images of the same eye with the same measurement method) are evaluated. For both intra and interscan reproducibilities, the intraclass correlation coefficient (ICC) and the coefficient of variation (CV) between measurements are computed. An ICC close to 1 indicates a high similarity between the measurements and a CV less than 1 indicates that there is a low variability in the measurements in relation to the mean of the measurements.

Scatterplots are used to show the relationship of each measurement such as examiner 1, examiner 2 and mKSM method.

Paired t-test and Bland-Altman plots are used to compare the differences between the measurements of the FAZ area among both examiners and mKSM method and the three plexuses. Statistical significance was defined as p<0.05.

## RESULTS

29 eyes of 25 healthy volunteers were included in the analysis (Table 1). None of the HS-OCTA images taken were excluded, although not all of them had good quality. From 174 images, KSM misidenfied 24 (14%) and overmeasured 30 (17%), while our method (mKSM) successfully measured all the images, showing that mKSM can be used for real-life images.

**Table 1.** Sample description.

| **Table 1. Sample description** |  |
| --- | --- |
| **SUBJECTS** |  |
| Age (years-old) | 36,6 (SD 11,8) |
| Caucasian (n) | 25 (100%) |
| Women (n) | 16 (64%) |
| Smoking history | 8 (32%) |
| **EYES** |  |
| Best-corrected visual acuity (Snellen) | 20/20±0 |
| Right eyes (n) | 21 (72,4%) |
| Phakic (n) | 28 (96,6%) |
| Axial length (mm) | 23,62±0,82 |
| *Abbreviations: Standard deviation (SD); number (n)* |  |

### Intra and Interscan Reproducibility of the FAZ Area

Table 2 shows the intra and interscan reproducibility of the FAZ area for both examiners and mKSM for the plexus SVP, ICP and DCP separately. Globally, for manual measurements, intrascan intraclass coefficient ranged from 0,948 to 0,993. For mKSM method this coefficient was 1, which means that mKSM always gives the same result for the same OCTA image.

**Table 2.**
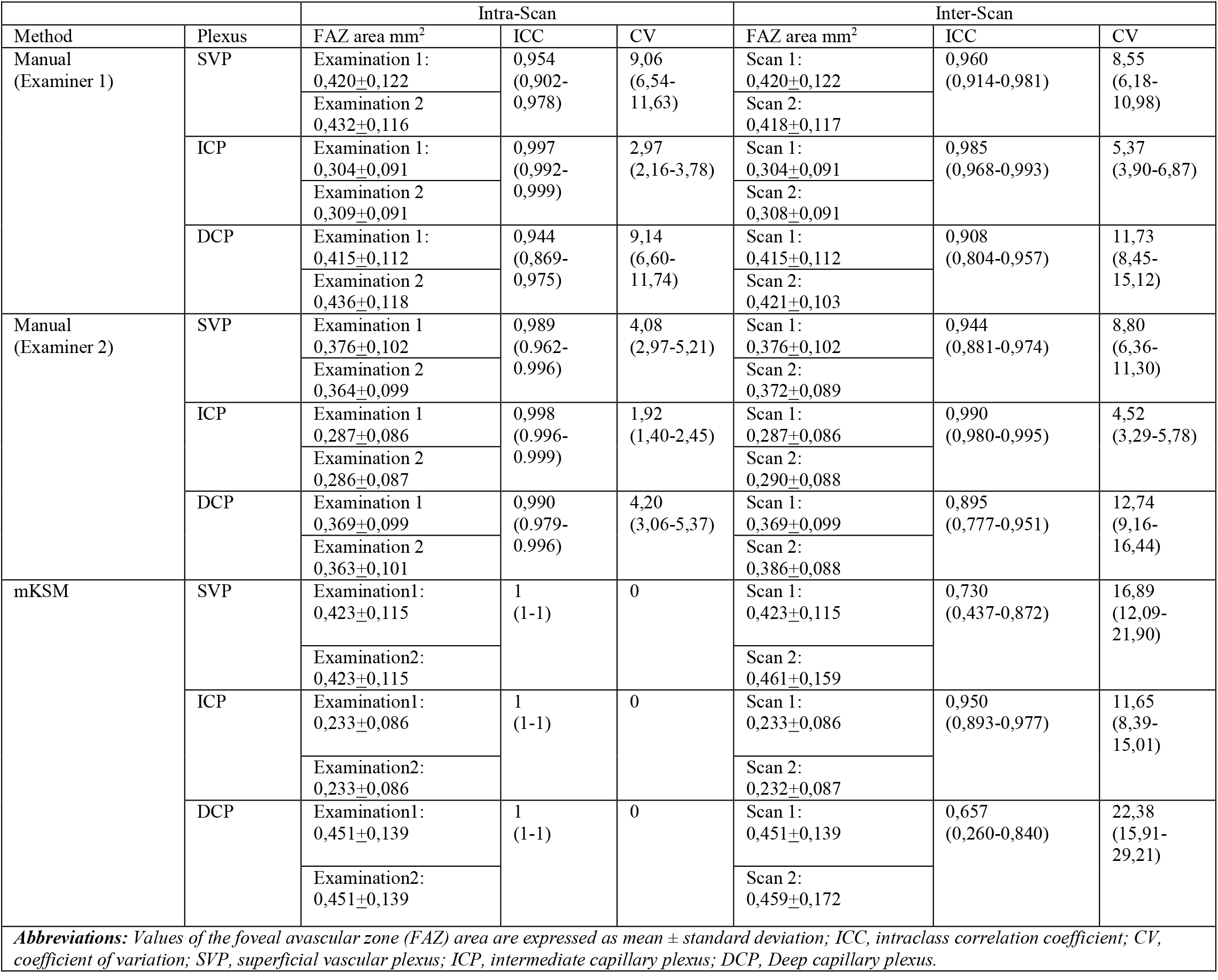
Intra and Interscan Reproducibility of Foveal Avascular Zone Area.

For manual measurements, the interscan intraclass coefficients ranged from 0,925 to 0,973. For mKSM method it ranged from 0,770 to 0,902. These results imply that for both human measurements and mKSM ones, the measured FAZ area for different images of the same eye are different, being the human measurements more consistent.

### Differences among the measurements

Figures 2, 3 and 4 show the differences among measurements between both human examiners (1 and 2) and mKSM for SVP, ICP and DCP FAZ areas. The mean difference and the 95% limits of agreement for each comparison are shown in Bland-Altman plots for every plexus. They show that the difference between human examiners is smaller than between human examiners and mKSM. Nevertheless, the scatterplots show a strong correlation between human and automatic measurements.

**Figure 3.**
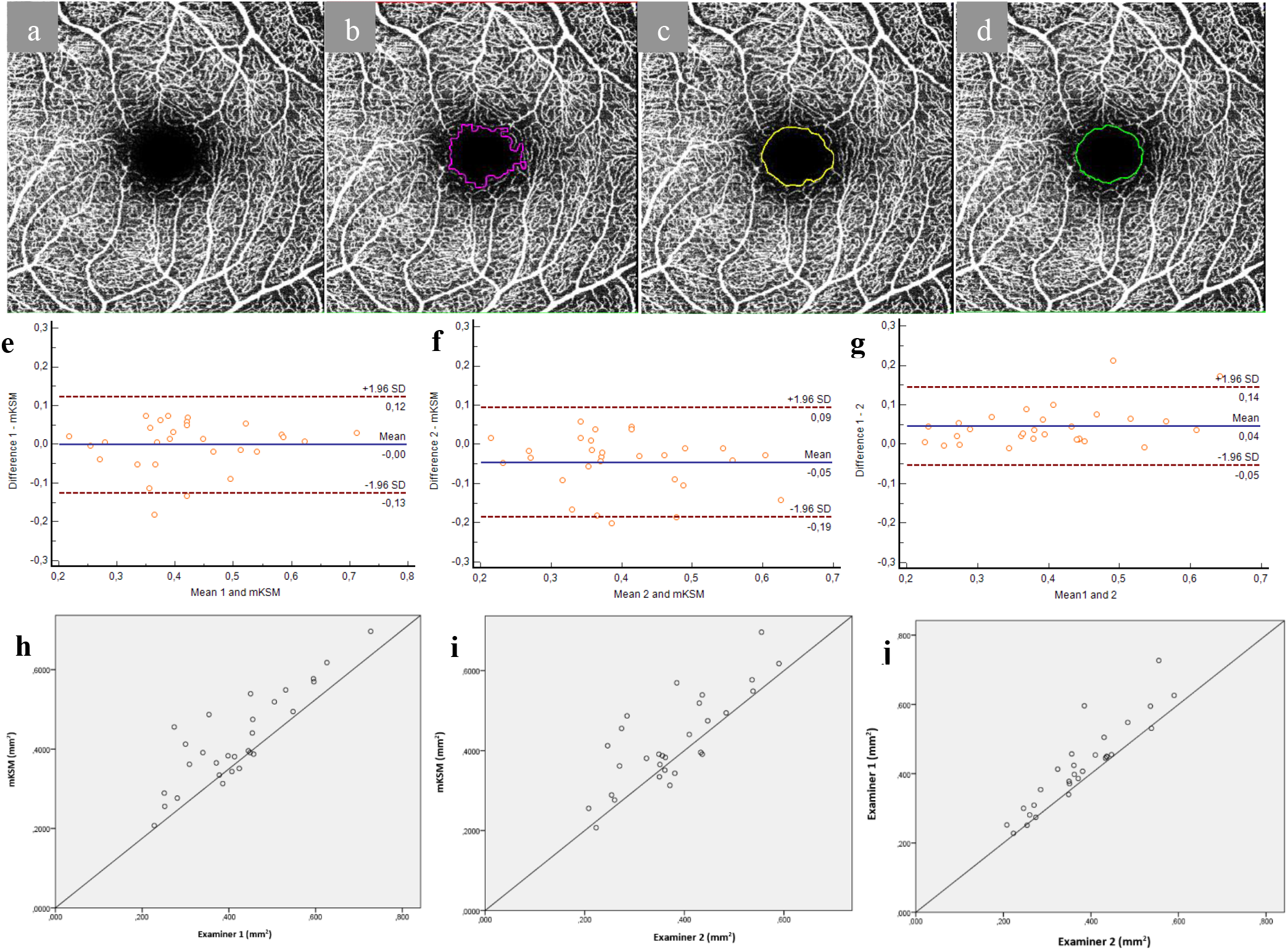
Optical coherence tomography angiography scan (a) showing the superficial vascular plexus. Foveal avascular zone area outlined by mKSM (b), examiner 1 (c) and examiner 2 (d). Below, Bland-Altman plots (e-g) and scatterplots showing the linear agreement (h-j) of the superficial vascular plexus foveal avascular zone measured by two examiners (1 and 2) and mKSM.

**Figure 4.**
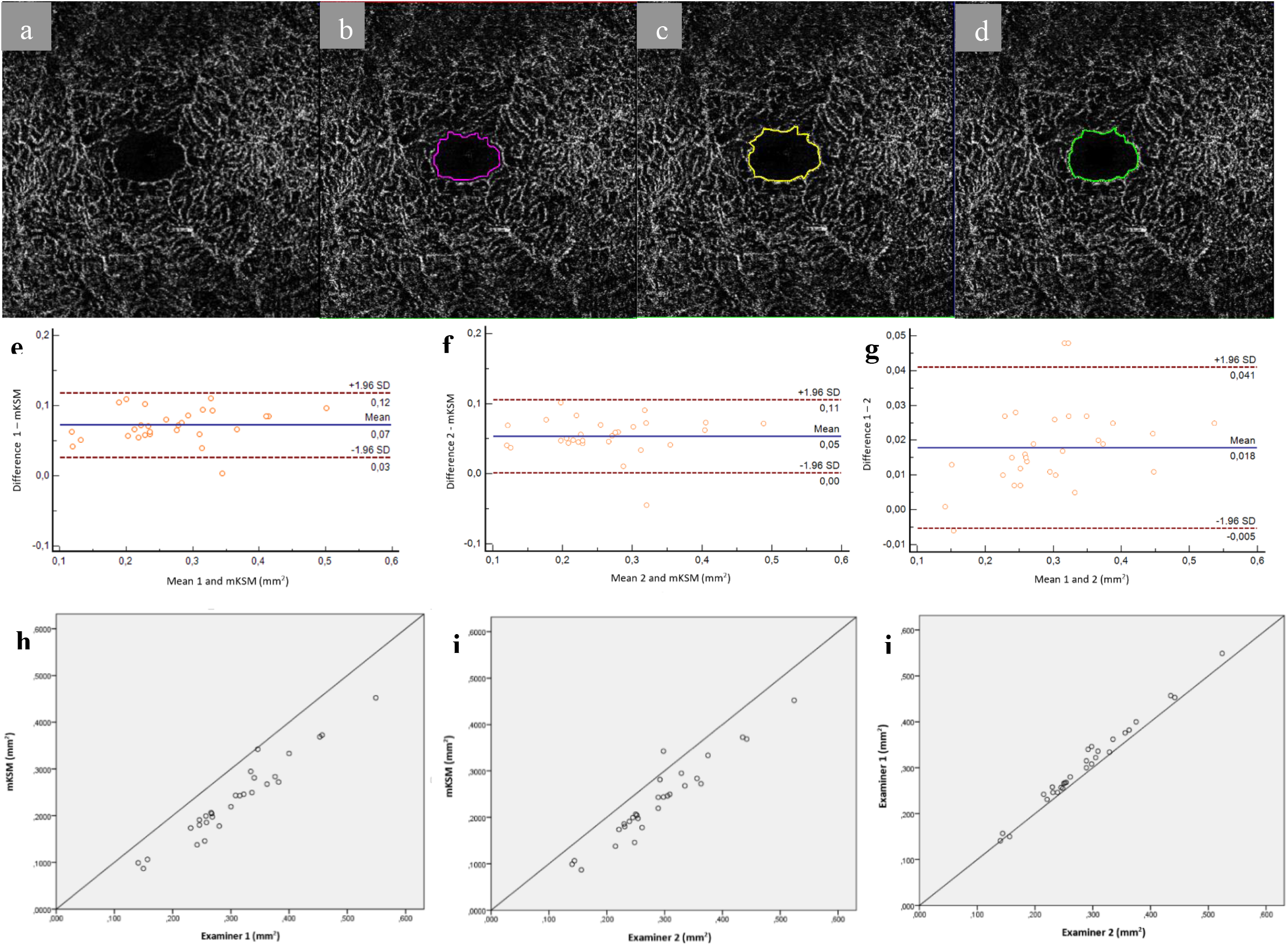
Optical coherence tomography angiography scan (a) showing the intermediate capillary plexus. Foveal avascular zone area outlined by mKSM (b), examiner 1 (c) and examiner 2 (d). Below, Bland-Altman plots (e-g) and scatterplots showing linear agreement (h-j) of the intermediate capillary plexus foveal avascular zone measured by two examiners (1 and 2) and mKSM.

**Figure 5.**
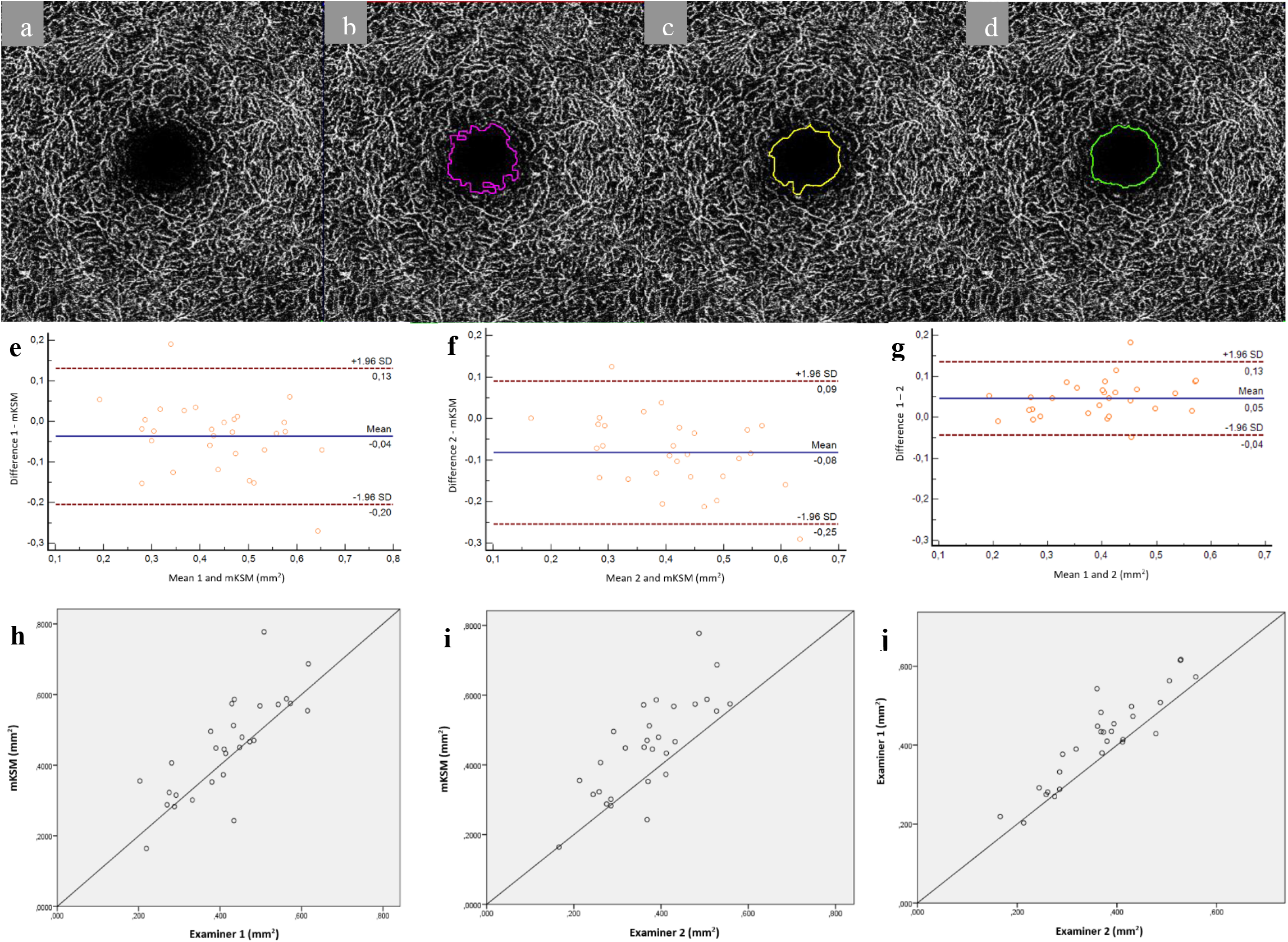
Optical coherence tomography angiography scan (a) showing the deep capillary plexus. Foveal avascular zone area outlined by mKSM (b), examiner 1 (c) and examiner 2 (d). Below, Bland-Altman plots (e-g) and scatterplots showing linear agreement (h-j) of the deep capillary plexus foveal avascular zone measured by two examiners (1 and 2) and mKSM.

The results of both human measurements and mKSM in ICP are the most consistent. However, the FAZ area measurement in ICP is systematically smaller for mKSM. This error could be palliated by a specific post-processed for ICP to force similar results to the human examiners. In this study, the processing of OCTA images has been the same for the three plexuses to make the processing plexus independent.

The results of the paired t-tests between the FAZ areas among the three methods show that the human measures of both examiners cannot be considered to belong to the same distribution (p=0,173 for examiner 1 vs mKSM; p=0,010 for examiner 2 vs mKSM; and p=0,000 for examiner 1 vs 2). This suggests that statistical hypothesis testing is not a suitable tool for assessing the quality of the measures of mKSM.

## DISCUSSION

Currently there are several studies in the literature about the FAZ area measurement in healthy eyes. Each study used a different device for the OCTA acquisition. Besides, some of them compared the results from different devices, concluding that the measurements of the FAZ area from different devices cannot be used interchangeably (28)(29)(30)(31). Moreover, HS-OCTA measures have been found to be systematically larger than Optovue RTVue XR Avanti(31).

The FAZ areas from all the OCTA devices can be measured manually, but only two of them have an embedded algorithm for the automated FAZ area measurement: Optovue RTVue XR Avanti (Optovue, Inc, Fremont, CA)(21) and Zeiss Cirrus HD-OCT 5000 with AngioPlex software (Carl Zeiss Meditec, Dublin, CA)(22). The interobserver agreement between the manual FAZ area measurements is good but it is poor when manual and automated FAZ area measurements are compared (Lin et al and LaSpina et al) (21)(22).

HS-OCTA (Software Version 6.12.1, Heidelberg Engineering, Heidelberg, Germany) does not have embedded software that automatically measures the FAZ area(24)(32) (33)(34). Therefore, we developed and evaluated an automated method to measure the FAZ area that relies on the processing tool angioTool and the scientific image processing tool ImageJ. This method is referred to as mKSM, and it is an evolution of the original KSM aproach.

The use of the mKSM method for the measurement of the FAZ area implies more reliability, reproducibility and speed in measurements compared to human manual measurements. While a human examiner takes about 1 minute to measure the FAZ area manually and its value changes with each measurement, the mKSM takes less than 3 seconds and its value is always the same for the same OCTA image.

Moreover, the mKSM is tolerant to images that are not perfectly centered or with artefacts, while the original KSM (27) only used images perfectly centered and without artefacts. This tolerance makes mKSM usable for images from the real world that could be acquired in a regular clinic.

Most of the studies only measure SVP(22), DCP(32) or a measurement that includes the 3 plexuses altogether(35) due to the believe that it allows to have less variability in the measures. However, in this study the three plexuses were measured separately: SVP, ICP, DCP. The mKSM processing is plexus independent, besides the best results are obtained in ICP.

Given that the FAZ area measurements among different devices are not interchangeably, the measurements of the current study can only be compared with other HS-OCTA studies where the FAZ area of healthy patients was measured (table 3). Only Lupidi et al. (32) used a semiautomated method to measure the FAZ area, while the others measured it manually (24)(33)(34). The mean SVP, ICP and DCP FAZ area results of the current study are similar to Hosari et al. (24) and somewhat larger than Pilotto et al. (33). The results of the current study are not comparable to Lupidi et al. (32) or Mihailovic et al. (34) because in the first one they used a prototype for the acquisition of the images and in the second one the images acquired had a worse resolution.

**Table 3.** Comparison of Heidelberg Spectralis studies.

| **Table 3. Comparison of Heidelberg Spectralis studies** |  |  |  |  |  |  |
| --- | --- | --- | --- | --- | --- | --- |
| *STUDY* (year) | IMAGING METHOD | NUMBER OF EYES | PATIENTS STATUS | PATIENTS AGE (years) | MEASUREMENT METHOD | MEAN FAZ AREA ± SD (mm<sup>2</sup>) |
| Current study (2020) | Heidelberg Spectralis (Lateral resolution: 5,7μm/pixel) | 29 | Healthy | 36,6±11,8 | Automated<br>Manual<br>Manual<br>Automated<br>Manual<br>Manual<br>Automated<br>Manual<br>Manual | SVP<br>0,423±0,115<br>0,420±0,122<br>0,376±0,102<br>ICP<br>0,233±0,0860,<br>0,304±0,091<br>0,287±0,086<br>DCP<br>0,451±0,139<br>0,415±0,112<br>0,369±0,099 |
| Hosari et al (2019) | Heidelberg Spectralis (Lateral resolution: 5,7μm/pixel) | 23 | Healthy | 26.8 ± 6 | Manual | SVP<br>0,42±0,23<br>ICP<br>0,28±0,1<br>DCP<br>0,44±0,12 |
| Lupidi et al (2016) | Prototype based on Heidelberg Spectralis (Lateral resolution: 6μm/pixel) | 47 | Healthy | 39 (19 –66) | Semiautomated | SVP<br>0.28 ± 0.11<br>DCP<br>0.30 ± 0.10 |
| Mihailovic et al (2018) | Heidelberg Spectralis (Lateral resolution: 14μm/pixel) | 24 | Healthy | 30.58± 12.11 | Manual | SVP<br>0.329± 0.095<br>DCP<br>0.335±0.093 |
| Pilotto et al (2018) | Heidelberg Spectralis (Lateral resolution: 6μm/pixel) | 33 | Healthy | 26 ± 3 | Manual | SVP<br>0.30 ± 0.08<br>DCP<br>0.35 ± 0.08 |
| **Abbreviations:** Values of the foveal avascular zone (FAZ) area are expressed as mean ± standard deviation; SVP, superficial vascular plexus; ICP, intermediate capillary plexus; DCP, Deep capillary plexus. | | | | | | |

In the current study, there are some limitations. On the one hand, the proband group is not very large, and the patients are young and healthy, so the results may not be extrapolable for older patients or with ocular pathologies, which should be needed for additional studies. On the other hand, only 2,9×2,9 mm scans of HS-OCTA were used, so our results could be not applicable for wider scans or OCTA images acquired with other devices.

Note that this study evaluates the proposed method in all OCTA images taken, without excluding bad quality images or images with discrepancies in the foveal area measures by different specialists. These results are expected to be worse quantitatively than other studies where the image sample has been biased in any of the previously mentioned directions.

In conclusion, automated determination of the FAZ area in HS-OCTA is feasible and less human-dependent by mKSM even if the image is not perfectly centered or has artefacts. There is variability among manual FAZ measurements by humans and among manual measurements and mKSM, although this variability is smaller in ICP FAZ area. mKSM method could contribute to our understanding of the three vascular plexuses, and could be used as a basis for future FAZ area processing.

## Data Availability

The current study is a cross-sectional study

## ACKNOWLEDGMENTS

This research received no specific grant from any funding agency in the public, commercial, or not-for-profit sectors.

## REFERENCES

1. Werner JU, Böhm F, Lang GE, Dreyhaupt J, Lang GK, Enders C. Comparison of foveal avascular zone between optical coherence tomography angiography and fluorescein angiography in patients with retinal vein occlusion. PLoS One. 2019 Jun 1;14(6).

2. Mochi T, Anegondi N, Girish M, Jayadev C, Roy AS. Quantitative comparison between optical coherence tomography angiography and fundus fluorescein angiography images: Effect of vessel enhancement. Ophthalmic Surg Lasers Imaging Retin. 2018 Nov 1;49(11):E175–81.

3. Soares M, Neves C, Marques IP, Pires I, Schwartz C, Costa MÂ, et al. Comparison of diabetic retinopathy classification using fluorescein angiography and optical coherence tomography angiography. Br J Ophthalmol. 2017 Jan 1;101(1):62–8.

4. JM G, TT L, RN L, AT R, DL I, M A. Diabetic Macular Ischemia Diagnosis: Comparison Between Optical Coherence Tomography Angiography and Fluorescein Angiography. J Ophthalmol. 2016;2016.

5. Spaide RF, Fujimoto JG, Waheed NK, Sadda SR, Staurenghi G. Optical coherence tomography angiography. Vol. 64, Progress in Retinal and Eye Research. Elsevier Ltd; 2018. p. 1–55.

6. Lavia C, Bonnin S, Maule M, Erginay A, Tadayoni R, Gaudric A. Vessel density of superficial, intermediate, and deep capillary plexuses using optical coherence tomography angiography. Retina. 2019 Feb;39(2):247–58.

7. Gómez-Ulla F, Cutrin P, Santos P, Fernandez M, Abraldes M, Abalo-Lojo JM, et al. Age and gender influence on foveal avascular zone in healthy eyes. Exp Eye Res. 2019 Dec 1;189:107856.

8. Ghassemi F, Mirshahi R, Bazvand F, Fadakar K, Faghihi H, Sabour S. The quantitative measurements of foveal avascular zone using optical coherence tomography angiography in normal volunteers. J Curr Ophthalmol. 2017 Dec 1;29(4):293–9.

9. Zhou Y, Zhou M, Gao M, Liu H, Sun X. Factors Affecting the Foveal Avascular Zone Area in Healthy Eyes among Young Chinese Adults. Biomed Res Int. 2020;2020.

10. Yu J, Jiang C, Wang X, Zhu L, Gu R, Xu H, et al. Macular perfusion in healthy chinese: An optical coherence tomography angiogram study. Investig Ophthalmol Vis Sci. 2015;56(5):3212–7.

11. Tan CS, Lim LW, Chow VS, Chay IW, Tan S, Cheong KX, et al. Optical coherence tomography angiography evaluation of the parafoveal vasculature and its relationship with ocular factors. Investig Ophthalmol Vis Sci. 2016 Jul 1;57(9):OCT224–34.

12. Fujiwara A, Morizane Y, Hosokawa M, Kimura S, Shiode Y, Hirano M, et al. Factors affecting foveal avascular zone in healthy eyes: An examination using swept-source optical coherence tomography angiography. PLoS One. 2017 Nov 1;12(11).

13. Wylęgała A, Wang L, Zhang S, Liu Z, Teper S, Wylęgała E. Comparison of foveal avascular zone and retinal vascular density in healthy Chinese and Caucasian adults. Acta Ophthalmol. 2020 Jun 26;98(4):e464–9.

14. Giocanti-Aurégan A, Gazeau G, Hrarat L, Lévy V, Amari F, Bodaghi B, et al. Ethnic differences in normal retinal capillary density and foveal avascular zone measurements. Int Ophthalmol. 2020 Nov 1;40(11):3043–8.

15. Tang FY, Ng DS, Lam A, Luk F, Wong R, Chan C, et al. Determinants of Quantitative Optical Coherence Tomography Angiography Metrics in Patients with Diabetes. Sci Rep. 2017 Dec 31;7(1):2575.

16. Koulisis N, Kim AY, Chu Z, Shahidzadeh A, Burkemper B, De Koo LCO, et al. Quantitative microvascular analysis of retinal venous occlusions by spectral domain optical coherence tomography angiography. PLoS One. 2017 Apr 1;12(4).

17. Kwon J, Choi J, Shin JW, Lee J, Kook MS. Alterations of the foveal avascular zone measured by optical coherence tomography angiography in glaucoma patients with central visual field defects. Investig Ophthalmol Vis Sci. 2017 Mar 1;58(3):1637–45.

18. Yu DY, Cringle SJ, Su EN. Intraretinal oxygen distribution in the monkey retina and the response to systemic hyperoxia. Investig Ophthalmol Vis Sci. 2005 Dec;46(12):4728– 33.

19. Gómez-Ulla F, Cutrin P, Santos P, Fernandez M, Abraldes M, Abalo-Lojo JM, et al. Age and gender influence on foveal avascular zone in healthy eyes. Exp Eye Res. 2019 Dec 1;189.

20. Zheng Y, Gandhi JS, Stangos AN, Campa C, Broadbent DM, Harding SP. Automated segmentation of foveal avascular zone in fundus fluorescein angiography. Investig Ophthalmol Vis Sci. 2010 Jul;51(7):3653–9.

21. La Spina C, Carnevali A, Marchese A, Querques G, Bandello F. Reproducibility and reliability of optical coherence tomography angiography for foveal avascular zone evaluation and measurement in different settings. Retina. 2017 Sep 1;37(9):1636–41.

22. Lin A, Fang D, Li C, Cheung CY, Chen H. Reliability of foveal avascular zone metrics automatically measured by Cirrus optical coherence tomography angiography in healthy subjects. Int Ophthalmol. 2020 Mar 1;40(3):763–73.

23. Corvi F, Pellegrini M, Erba S, Cozzi M, Staurenghi G, Giani A. Reproducibility of Vessel Density, Fractal Dimension, and Foveal Avascular Zone Using 7 Different Optical Coherence Tomography Angiography Devices. Am J Ophthalmol. 2018 Feb 1;186:25–31.

24. Hosari S, Hohberger B, Theelke L, Sari H, Lucio M, Mardin CY. OCT Angiography: Measurement of Retinal Macular Microvasculature with Spectralis II OCT Angiography-Reliability and Reproducibility. Ophthalmologica. 2020 Jan 1;243(1):75– 84.

25. Campbell JP, Zhang M, Hwang TS, Bailey ST, Wilson DJ, Jia Y, et al. Detailed Vascular Anatomy of the Human Retina by Projection-Resolved Optical Coherence Tomography Angiography. Sci Rep. 2017 Feb 10;7.

26. Zudaire E, Gambardella L, Kurcz C VS. A Computational Tool for Quantitative Analysis of Vascular Networks. PLoS One. 2011;6(11):e27385.

27. Ishii H, Shoji T, Yoshikawa Y, Kanno J, Ibuki H, Shinoda K. Automated measurement of the foveal avascular zone in swept-source optical coherence tomography angiography images. Transl Vis Sci Technol. 2019 May 1;8(3).

28. Corvi F, Pellegrini M, Erba S, Cozzi M, Staurenghi G, Giani A. Reproducibility of Vessel Density, Fractal Dimension, and Foveal Avascular Zone Using 7 Different Optical Coherence Tomography Angiography Devices. Am J Ophthalmol. 2018 Feb 1;186:25–31.

29. Magrath GN, Say EAT, Sioufi K, Ferenczy S, Samara WA, Shields CL. Variability in foveal avascular zone and capillary density using optical coherence tomography angiography machines in healthy eyes. Retina. 2017 Nov 1;37(11):2102–11.

30. Dave PA, Dansingani KK, Jabeen A, Jabeen A, Hasnat Ali M, Vupparaboina KK, et al. Comparative Evaluation of Foveal Avascular Zone on Two Optical Coherence Tomography Angiography Devices. Optom Vis Sci. 2018 Jul 1;95(7):602–7.

31. Anvari P, Najafi A, Mirshahi R, Sardarinia M, Ashrafkhorasani M, Kazemi P, et al. Superficial and deep foveal avascular zone area measurement in healthy subjects using two different spectral domain optical coherence tomography angiography devices. J Ophthalmic Vis Res. 2020 Oct 1;15(4):517–23.

32. Lupidi M, Coscas F, Cagini C, Fiore T, Spaccini E, Fruttini D, et al. Automated Quantitative Analysis of Retinal Microvasculature in Normal Eyes on Optical Coherence Tomography Angiography. Am J Ophthalmol. 2016 Sep;169:9–23.

33. Pilotto E, Frizziero L, Crepaldi A, Della Dora E, Deganello D, Longhin E, et al. Repeatability and Reproducibility of Foveal Avascular Zone Area Measurement on Normal Eyes by Different Optical Coherence Tomography Angiography Instruments. Ophthalmic Res. 2018 Feb 8

34. Mihailovic N, Brand C, Lahme L, Schubert F, Bormann E, Eter N, et al. Repeatability, reproducibility and agreement of foveal avascular zone measurements using three different optical coherence tomography angiography devices. Madigan M, editor. PLoS One. 2018 Oct 18;13(10):e0206045.

35. Garrity ST, Iafe NA, Phasukkijwatana N, Chen X, Sarraf D. Quantitative analysis of three distinct retinal capillary plexuses in healthy eyes using optical coherence tomography angiography. Investig Ophthalmol Vis Sci. 2017 Oct 1;58(12):5548–55.

